# Evaluating Frailty Index Integrity: Insights from an International Network Study

**DOI:** 10.64898/2026.01.12.26343698

**Authors:** Robert Cavanaugh, Chelsea Wong, Louisa H. Smith, Maytal Bivas-Benita, Pinchas Akiva, Tal El-Hay, Ariela R. Orkaby, Chen Yanover, Brianne Olivieri-Mui

**Affiliations:** MGH Institute of Health Professions, Boston, MA; The Roux Institute, Northeastern University, Portland, ME; Northeastern University, Boston, MA; Beth Israel Deaconess Medical Center, Boston; KI Research Institute, Kfar Malal, Israel; New England GRECC (Geriatric Research, Education and Clinical Center), VA Boston Healthcare System, Boston, MA; Division of Aging, Brigham & Women’s Hospital, Harvard Medical School, Boston, MA; The Marcus Institute for Aging Research, Hebrew Senior Life, Harvard Medical School, Roslindale, MA

## Abstract

Deficit-accumulation Frailty Indices (FIs) have rapidly been integrated into health systems worldwide to quantify the state of reduced physiologic reserve to recover from a health insult in the aging population. Common data models (CDMs) have further enabled international, interinstitutional applications of FIs. However, the validity of such applications is unknown.

We conducted an international network study comparing estimates of frailty from two electronic health record (EHR)-based FIs: United States (US)-based Veterans Affairs FI (VAFI) and United Kingdom (UK)-based electronic FI (eFI) across 5 research and clinical databases. US: *All of Us* (n=159,721), IQVIA Pharmetrics+ (n=5,099,557); UK: IMRD-THIN (n=3,036,003), IMRD-EMIS (n=832,455), and BioBank (n=207,202).

In US databases, VAFI identified higher proportions of frailty ([VAFI; eFI] *All of US*: 10.3%; 2.5%, Pharmetrics+: 9.6%; 2%) while in UK databases, the eFI identified higher proportions (IMRD-THIN: <0.003%; 0.08%, IQVIA-EMIS: 0.09%; 0.4%, UK BioBank: 0.03%; 0.1%). Additional manipulations (alternative lookback periods, FI variations) were examined.

FIs are likely dependent on their development context, such as local coding behaviors and incentives, limiting their external validity despite CDM harmonization. We suggest caution in the application of FIs outside of their development context and recommend further instrument development before more widespread use.

## Introduction

Frailty is a state of physiologic vulnerability that identifies older adults at risk of poor outcomes such as cardiovascular events, falls, hospitalization, and death (1,2). Automated frailty indices (FIs) are derived from the gold standard Comprehensive Geriatric Assessment, which includes multiple domains of health, including cognition, morbidity, physical function, and mental health, to measure frailty (3–5). The benefit of the FI approach has been widely validated and is increasingly being adapted for health systems to guide population health and clinical decision making (1,6).

Distinct FIs have been developed using observational and administrative health data such as the hospital frailty risk score (HFRS) developed in hospitalized patients aged 75 and older in the UK (7), the claims-based frailty index (CFI) developed in US Medicare claims data for adults aged 65 and older (8), the Veterans’ Affairs frailty index (VAFI), derived from electronic health record data from US Veterans aged 45 and older (9,10), and the electronic frailty index (eFI) developed from UK primary care data in adults aged 65 and older (11). Each was validated for specific geographic or demographic populations. Although crosswalks have been developed to compare different clinical tools to measure frailty (12,13), direct comparison and harmonization of different automated FIs across diverse international cohorts has not been attempted at scale.

The growing use of common data models (CDMs) such as the Observational Medical Outcomes Partnership (OMOP) CDM facilitates the concurrent analysis of data across institutions and provides the infrastructure necessary to calculate and compare FIs across international databases (14). CDMs can help overcome challenges associated with international comparisons of frailty to date: data heterogeneity and inconsistencies in how data are captured. This allows for more robust analyses and data interoperability, facilitating query sharing across multiple organizations, and maintaining privacy of patient-level data (15). To this end, frailty indices (e.g., HFRS) are integrated into existing CDM software so that they can be easily calculated across databases with many additional cohort characteristics (16).

However, while CDMs enable calculation of frailty across international databases, they do not ensure that FIs are valid outside of the contexts in which they were developed. In such cases, adaptations are required to adjust existing FIs to local data. For example, the UK-based eFI was previously adapted for healthcare centers in Barcelona, resulting in the eFRAGICAP frailty index (17). Without adaptation, applications outside of the original setting run the risk of misclassification. As such, we sought to examine, first, the differences in frailty prevalence for two FIs across two US and three UK EHR databases formatted for the OMOP CDM and, second, to demonstrate fidelity of the FIs within these databases.

## Methods

### Study design

This was a multinational retrospective cohort study using routinely collected healthcare data from both the US and UK, standardized to the OMOP CDM (18).

### Data sources

We examined frailty across 5 data sources:

IQVIA PharMetrics+® for Academics (2017–2022) includes US commercial claims for inpatient and outpatient healthcare and prescriptions. The source population includes people enrolled with private health insurers, select managed care plans, and supplements for Medicaid and Medicare. These data do not allow for following people switching insurers, limiting longitudinal analysis, and include only age and binary sex demographic data. PharMetrics+ is licensed to Northeastern University, and use of the data received a non-human subjects research determination from the Northeastern University Internal Review Board (IRB).

*All of Us* (AoU) Research Program data (V7; accessed July 2025) represents a convenience sample of more than 400,000 participants across the US, with an emphasis on oversampling groups historically underrepresented in research (19). These data include self-report surveys on personal demographics, family and personal health and lifestyle, data from AoU admission physical exams (e.g., height, weight, blood pressure), as well as EHR from contributing regional health centers, federally qualified health centers, Veterans Affairs medical centers, and genomic data from biospecimens. All experimental protocols and data collection involving human participants are covered by the Ethics Committee/IRB of the *All of Us* Research Program and the IRB at Northeastern University (#221016).

IQVIA Medical Research Data (IMRD) contains longitudinal non-identified patient EHR collected from two different UK General Practitioner (GP) clinical systems (20):

i) IMRD-THIN (The Health Improvement Network) (version: 2022-09, 17M individuals) is a Cegedim database where UK practices (in England, Wales, Scotland, and Northern Ireland) are recruited to contribute longitudinal patient data
ii) IMRD-EMIS contains data from practices using the EMIS practice management software in England (version: 2022-12, 5.4M individuals).

The use of IMRD for research has been approved by the NHS Health Research Authority (NHS Research Ethics Committee ref 18/LO/0441) for medical and public health research; this study received Scientific Review Committee (SRC) approval Ref 23SRC015. The validity of these transformed data has been demonstrated (21).

The UK Biobank (UKBB) is an ongoing prospective cohort study of over 500,000 participants, residents of England, Scotland, and Wales, recruited in 2006-2010 between ages 40-69 years (22,23). Participants completed a set of questionnaires (e.g., diet and well-being), underwent a brief interview, and had their physical measurements and biological samples taken. Self-report data have been linked to EHRs for most participants. UK Biobank has received ethical approval from the UK National Health Service’s National Research Ethics Service (reference 11/ NW/0382), and this work is part of project number 58770. As the research is based on the UK Biobank data, informed consent and the Declaration of Helsinki were not relevant/necessary.

### Study Samples

The index date for entry into the study sample was established in two ways, depending on database type. For *All of Us* and the UKBB, we used the date 365 days after study enrollment to ensure sufficient lookback after entering each cohort. For PharMetrics+, IMRD-THIN, and IMRD-EMIS, a majority of participants are present in the data on the first date of data availability. Therefore, to minimize bias from selection and temporality, we selected a random visit date for each unique person-identifier to serve as the index date.

To be included in the study, the participants had to be > 40 years old at the index date and had to have at least one year of continuous observation prior to the index date in the US databases and three years in the UK databases. While frailty was estimated for 1-year lookback periods for all databases, three years was chosen as the inclusion criterion for the UK databases because UK practice standards do not include coding all conditions at each encounter (24). This criterion further allowed us to manipulate the U.K. lookback period length to 3 years in sensitivity analyses (noted below). We were unable to use a similar 3-year lookback in the US databases due to a lack of sufficient longitudinal data. Age >40 was used to be inclusive and was supported by the age ranges analyzed in a 2018 meta-analysis demonstrating the utility of an FI for mortality risk across ages from 40 to 80+ (25,26).

### Demographics

Demographic information was largely limited to age and sex for all databases due to the deidentified nature of the data. Because individuals > 80 years old were indicated as being 80 years old in several databases, we grouped age into 5-year bins starting at age 40 and ending at >= 80 years old.

### Frailty Indices

For each FI, deficits were considered present if the person had evidence of any of the relevant concept identifiers (i.e., the OMOP vocabulary equivalent of ICD, SNOMED, CPT codes) and absent if none were present during the lookback period. The frailty index of an individual was computed by dividing the number of present deficits by the maximum possible deficits.

#### Veterans Affairs Frailty Index (VAFI)

Construct validity for the VAFI has been demonstrated against other frailty assessments and used in non-veteran populations (27,28). This FI uses a list of International Classification of Disease version 10 (ICD-10), current procedural terminology (CPT), and healthcare common procedure coding (HCPCS) codes, available at the VA Boston GitHub (https://github.com/bostoninformatics/va_frailty_index), across 31 deficits. The ICD codes were mapped to Systematized Nomenclature of Medicine – Clinical Terms (SNOMED-CT) clinical terminology codes using OHDSI vocabulary relationships (18). CPT codes were mapped directly to OMOP concept IDs. Codes which did not map 1:1 to SNOMED-CT were independently manually reviewed by two geriatricians (CW and AO) to ensure that the mapped OMOP concept codes and their definitions were appropriate FI deficits. As a check of the accuracy of our transformation of the ICD and CPT VAFI codes to OMOP standard vocabularies, we compared the prevalence of each deficit for the VAFI based on the OMOP concept IDs to the prevalence based on ICD values from the source data (Supplement Figure 1). The excellent intraclass correlation (>0.98) ensured that the resulting OMOP FI code-list was in high agreement with the ICD source codes for the VAFI. Individual VAFI scores were computed by dividing the number of present deficits by 3. Categories of robust/fit (£0.1), pre-frail (>0.1–0.2), and frail (mild: >0.2–0.3, moderate: >0.3–0.4, severe: >0.4) were determined by published cut-points (29).

#### Electronic Frailty Index (eFI)

Proprietary SNOMED-CT codes for calculating eFI were determined by and provided to our study team by Andrew Clegg (30). The SNOMED-CT codes were manually mapped to OMOP standard concepts for analysis. The eFI can include laboratory measures not available in the US data, including hemoglobin estimation (anemia and hematinic deficiency), urine albumin or protein levels (chronic kidney disease), and Thyroid-stimulating hormone (TSH) levels (thyroid disease). Therefore, we did not include these measurements in our base comparison but did include them for the UK databases for sensitivity analyses. We also report polypharmacy across databases, estimated using methods described in Elhussein et al (31), because polypharmacy codes were not used consistently across databases. We defined polypharmacy as having a clinical finding of polypharmacy or prescriptions for at least 10 unique ingredients, excluding antibiotics identified as any drug in ATC J01, in a 1-year lookback. There are 36 possible deficits in the eFI, and the discrete eFI is typically categorized as robust/fit (£0.12), mild frailty (>0.12–0.24), and frail (moderate frailty >0.24–0.36, severe frailty >0.36) based on published cut-points (30). To match the VAFI, we reduced these categories to robust or fit (£0.12), mild frailty (pre-frail; >0.12–0.24), and frail (>0.24).

### Analysis

Extraction of frailty concepts from the OMOP CDM and calculation of prevalence of frailty and overall frailty categories was completed using a unified code base written in R (R Core Team) using the R package dbplyr (32), which translates R code into the appropriate SQL code for each database (Amazon Redshift, Google BigQuery, etc.). Analytical code and aggregated summary data are available at: https://github.com/rbcavanaugh/frailty-comparisons. Pharmetrics+, IMRD-EMIS, and IMRD-THIN are all IQVIA data products licensed to our institutions, and individual-level data is not publicly available. Data within the *All of Us* Research Program and the UK BioBank are available through the respective programs.

### Sensitivity Analyses

To minimize the chances that our choices of lookback period and exclusion of measurement data from the eFI in databases (where available) biased our results, we modified our estimates of frailty to test the robustness of the findings in the following ways: (1) We increased the lookback period in UK databases from one year to three years, based on feedback from clinicians that UK healthcare systems do not “carry forward” diagnoses from past visits to the extent that is done in the US. This might require looking back further to obtain comparable estimates of frailty between the healthcare systems. Only the eFI includes laboratory measures in estimating frailty, and these measurements were only available in UK databases and inconsistently in the *All of Us* program data, so initial estimates of frailty excluded measurement data across all databases. To ensure this did not unduly influence our findings, we re-estimated frailty using measurement data in the UK databases. We examined the four combinations of these sensitivity analyses (1 and 3-year lookback and with and without measurements in the UK data) in assessing the robustness of our findings. The effects of these sensitivity analyses are visualized in the interactive website reported in the results.

## Results

The final sample sizes across databases were: *All of Us* n=159,721, US IQVIA Pharmetrics+ n=5,099,557, UK IMRD-THIN n=3,036,003, UK IMRD-EMIS n=832,455, and UK BioBank n=207,202. The effects of inclusion criteria on sample sizes for each database are reported in Supplemental Table 1.

Cohort demographics and overall frailty prevalence are reported in Table 1. Additionally, we provide an interactive visualization of the differences in frailty prevalence across data sources and by sex for each frailty measure overall and within each frailty deficit category at https://roux-ohdsi.observablehq.cloud/interactive-frailty-indices/.

**Table 1.** Baseline characteristics of cohorts from each database.

|  | Pharmetrics+<br>(N=5,099,557) | All of Us<br>(N=159,721) | IMRD-EMIS<br>(N=832,455) | IMRD-THIN<br>(N=3,036,003) | UK BioBank<br>(N=207,202) |
| --- | --- | --- | --- | --- | --- |
| **Sex (at birth)** |  |  |  |  |  |
| Female | 2,735,536 (53.6%) | 97,658 (61.1%) | 418,408 (50.3%) | 1,598,166 (52.6%) | 114,384 (55.2%) |
| Male | 2,364,021 (46.4%) | 62,063 (38.9%) | 414,047 (49.7%) | 1,437,837 (47.4%) | 92,818 (44.8%) |
| **Age (years)** |  |  |  |  |  |
| [40,45) | 596,638 (11.7%) | 14,626 (9.2%) | 105,426 (12.7%) | 353,147 (11.6%) | 12,939 (6.2%) |
| [45,50) | 610,663 (12.0%) | 15,913 (10.0%) | 105,775 (12.7%) | 390,736 (12.9%) | 25,688 (12.4%) |
| [50,55) | 657,697 (12.9%) | 19,613 (12.3%) | 109,520 (13.2%) | 395,711 (13.0%) | 30,010 (14.5%) |
| [55,60) | 702,585 (13.8%) | 23,709 (14.8%) | 99,193 (11.9%) | 359,165 (11.8%) | 35,548 (17.2%) |
| [60,65) | 698,925 (13.7%) | 23,922 (15.0%) | 89,493 (10.8%) | 336,935 (11.1%) | 48,759 (23.5%) |
| [65,70) | 574,070 (11.3%) | 22,783 (14.3%) | 83,563 (10.0%) | 328,897 (10.8%) | 43,159 (20.8%) |
| [70,75) | 439,984 (8.6%) | 19,039 (11.9%) | 73,321 (8.8%) | 271,880 (9.0%) | 11,099 (5.4%) |
| [75,80) | 335,448 (6.6%) | 11,535 (7.2%) | 60,635 (7.3%) | 223,491 (7.4%) | 0 (0%) |
| [80,120] | 483,547 (9.5%) | 8,581 (5.4%) | 105,529 (12.7%) | 376,041 (12.4%) | 0 (0%) |
| **eFI Frailty Category** |  |  |  |  |  |
| frail | 103,721 (2.0%) | 5,333 (3.3%) | 3,886 (0.5%) | 3,097 (0.1%) | 283 (0.1%) |
| prefrail | 536,434 (10.5%) | 22,499 (14.1%) | 45,259 (5.4%) | 95,111 (3.1%) | 7,061 (3.4%) |
| robust | 4,459,402 (87.4%) | 131,889 (82.6%) | 783,310 (94.1%) | 2,937,795 (96.8%) | 199,858 (96.5%) |
| **VAFI Frailty Category** |  |  |  |  |  |
| frail | 497,290 (9.8%) | 19,034 (11.9%) | 753 (0.1%) | 129 (0.0%) | 69 (0.0%) |
| prefrail | 892,252 (17.5%) | 29,982 (18.8%) | 16,016 (1.9%) | 8,114 (0.3%) | 1,541 (0.7%) |
| robust | 3,710,015 (72.8%) | 110,705 (69.3%) | 815,686 (98.0%) | 3,027,760 (99.7%) | 205,592 (99.2%) |

*Note: Race/ethnicity is not included in Table 1 because these data are not available from the IQVIA products due to the deidentified nature of those data. UK BioBank does not collect race/ethnicity data. eFI frailty categories cut-points for robust (0–0.12), pre-frail (>0.12–0.24), and frail (>0.24); VAFI frailty category cut-points for robust (≤0.1), pre-frail (>0.1–0.2), and frail (>0.2)*

All UK and Pharmetrics+ databases were approximately half female, while the US *All of Us* database was >60% female (Table 1). A plurality of the Pharmetrics+ sample were between ages 50-65 (40.4%), while *All of Us* was aged 55-70 (44.1%), IMRD-EMIS was aged 40-55 (38.5%), and IMRD-THIN was aged 45-60 (37.7%); IMRD-EMIS was the youngest sample. The UKBB does not have data for ages past 75, and the largest proportion (61.5%) was between 55-70. The UKBB and All of Us were the oldest samples.

Within the US databases, the VAFI identified higher levels of frailty than the eFI (Pharmetrics+ VAFI 9.8% vs. eFI 2.0%; All of Us 11.9% vs. 3.3%). Within the UK databases, the relationship was reversed (IMRD-EMIS VAFI: 0.1% vs. eFI: 0.5%; IMRD-THIN <0.1% vs. 0.1%; UKBB <0.1% vs. 0.1%) (Table 1).

Using either FI, the frailest database overall was *All of Us,* which also had the highest proportion of female individuals and skewed towards older ages. The least frail database by either FI measure was IMRD-THIN, though only slightly less than UKBB. Despite skewing younger in age, IMRD-EMIS had the highest frailty among the UK databases. By sex, Figure 1 shows that females had slightly higher frailty and pre-frailty in the US databases but were indistinguishable from males in the UK databases for the VAFI. For the eFI, males and females were generally indistinguishable across frailty categories (Figure 1, panel B).

**Figure 1.**
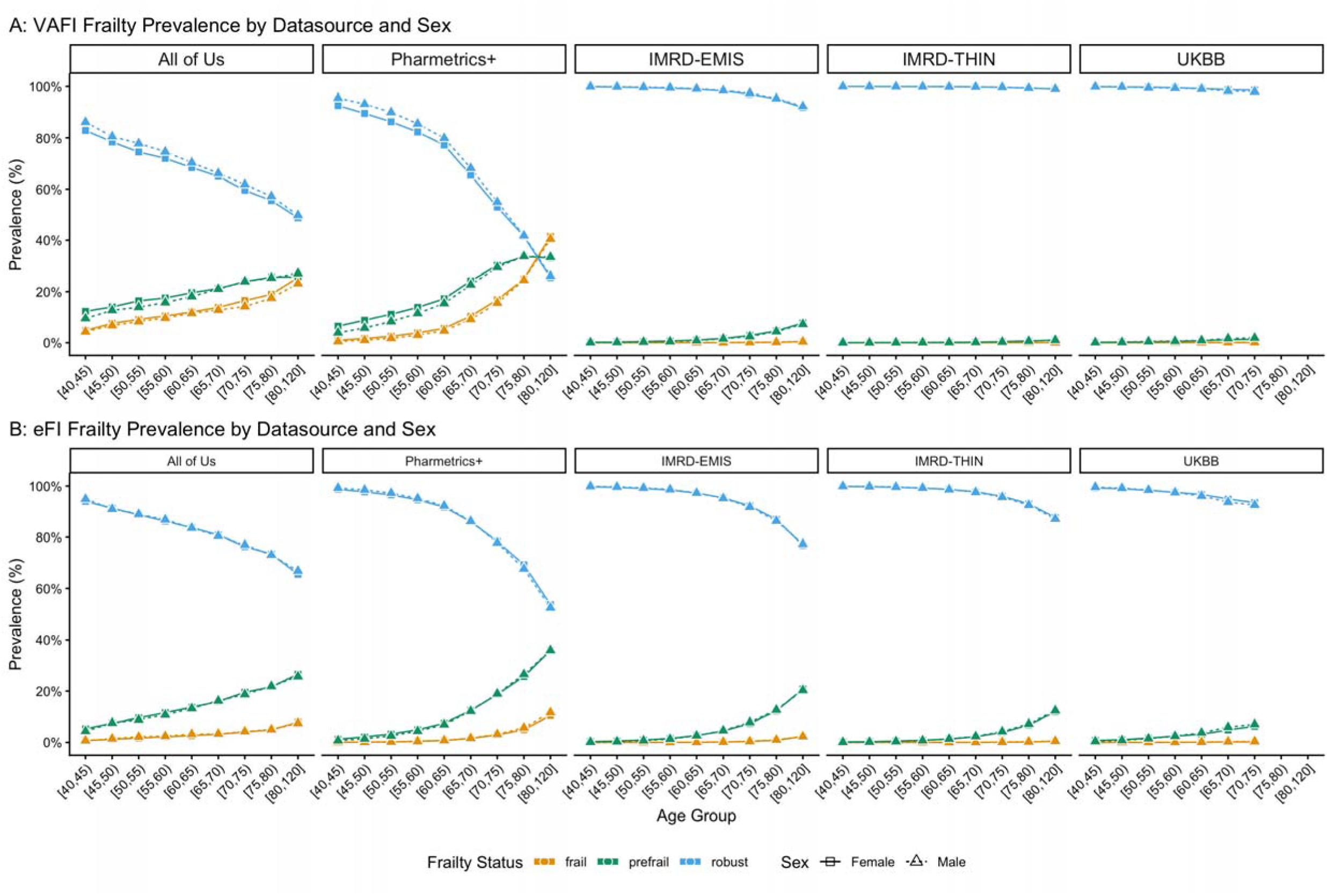
Panel A (top) VAFI Frailty Prevalence by Datasource and Sex, and Panel B (bottom) eFI Frailty Prevalence by Datasource and Sex, and Panel.

In our sensitivity analysis of the UK samples, which had three years of data, we found that using a three-year lookback period only modestly increased the proportion of the sample classified as frail across ages. The difference at the oldest age groups ranged from approximately 1 percentage point greater frailty in UKBB to 4.8 percentage points greater in IMRD-THIN and 8.6% greater frailty in IMRD-EMIS. When measurements were included in the eFI calculation for UK databases with a 3-year lookback, there was an additional reduction in robust classification. In the oldest age groups, IMRD-EMIS showed <3 Percentage points greater frailty across UK databases. Full results of the sensitivity analysis are reported in Supplemental Table 2.

## Discussion

Using a network study approach, we calculated 2 FIs across 2 US and 3 UK databases harmonized using the OMOP CDM. We aimed to demonstrate differences in frailty across international clinical settings and to understand the fidelity of FIs in data transformed to a CDM. On the surface, the US cohorts appeared more frail than the UK, but persistent variability between the FIs and across databases suggests the influence of exogenous factors. Specifically, the finding that VAFI demonstrated greater frailty in the US while the eFI demonstrated greater frailty in the UK suggests that frailty, as evidenced by these two FIs, may be more dependent on the nature of the source data, rather than the true prevalence of frailty in each population. In the following, we discuss potential reasons for these findings and suggest potential features of the source data that may have contributed to these results.

First, we found extreme variability in frailty prevalence in all databases from both the eFI and VAFI that we suspect is attributable to the data transformation process. For example, frailty prevalence tends to increase as age advances, but at nearly 40% in Pharmetrics+ and a maximum of 25% frail in *All of Us* by age 80+ using the VAFI, frailty was far more prevalent than expected. The eFI returned US results closer to expected in the oldest populations, around 10% frail (33). Where, in the UK, established estimates suggest frailty prevalence should be around 8%, varying by rurality and coastal proximity (34). At less than 5%, our results imply a much lower prevalence of frailty in the UK than other studies; the VAFI identified the least frailty, while the eFI identified the highest frailty at 3.25% in IMRD-EMIS. In contrast, the validation study for eFI, which used IMRD-THIN data in its original non-CDM format, demonstrated 43% robust, 37% pre-frail, and 20% frail (30). This discrepancy could be related to the use of a problem list in UK practice, which is otherwise excluded from the Extract, Transform, Load (OMOP CDM transformation) process.

The transformation of source data to a CDM-compatible vocabulary requires interpreting disparate health data into a common language and format. In this case, the OMOP CDM uses SNOMED-CT language. Therefore, the transformation is only as accurate as the ability to make a 1:1 translation of certain components of health records. However, there is often a one-to-many or many-to-one transformation of US-centric ICD-9 or ICD-10 codes to SNOMED-CT codes. Challenges with translation might explain discrepancies between our study and frailty originally reported for THIN during the validation study for the eFI (30). Moreover, the inclusion of laboratory measures in our sensitivity analyses did little to shift eFI frailty prevalence in the UK data towards the original study outcomes. Our results suggest that FIs like the eFI lack external validity when used with CDMs lacking such features as the problem list or a 1:1 translation of all data features.

Second, transformation to the OMOP CDM cannot overcome source data quality challenges such as missing data, misclassification, or misdiagnosis that stem from practice, billing, and cultural norms around healthcare utilization (35). In the UK, the NHS is a large publicly funded universal health care system (36); in the UK, the NHS is a large publicly funded universal health care system; all UK residents have compulsory membership that makes healthcare accessible and affordable. The US, however, relies on voluntary engagement with insurance coverage, where significant numbers of uninsured or underinsured individuals without consistent access to affordable healthcare in a landscape made up of a mix of fee-for-service and bundled payment models in a multi-payer system (37,38). Moreover, there is no single insurance system in which US residents are engaged because there are options such as Medicare, Medicaid, private insurance, or the insurance marketplaces for each state, depending on age, income, or employment. Therefore, in the current payment model, US providers are often incentivized to document all conditions at each encounter, assisted by EHR systems that often default to “carrying forward” diagnoses from past visits to the present, despite the existence of a problem list (39). In contrast, standard practice in the UK is to maintain and rely on a problem list for active conditions, precluding condition documentation at each visit (40). Therefore, it is likely that the discrepancies in the prevalence of frailty between US and UK data in a one-year lookback period were due to differences in healthcare documentation practices rather than true differences in condition prevalence and subsequent frailty prevalence.

That said, documentation is also dependent on who is seeking healthcare. The sample captured by each dataset represented an array of convenience samples from both countries. Even though the UKBB had a sampling frame, their efforts at a representative sample were diminished by a 5.5% response rate (23). The *All of Us* research program in the US seeks to oversample individuals from groups underrepresented in biomedical research, but does not have a sampling frame (41). Subsequently, both UKBB and *All of Us* display volunteer bias: volunteers are generally less frail and more likely to be female (42,43). In comparing sexes, we saw little difference in levels of frailty, which is contrary to known statistics that show women experience frailty more often than men as they age (44). Additionally, IMRD-EMIS is a medical record program widely used in the UK, and IMRD-THIN is a research-oriented dataset from providers that are all trained for data collection and entry, who volunteer to contribute their practices’ data and to use the Vision medical record program (45–47). Individuals in IMRD-THIN were more often robust in our study and are considered more broadly representative of the UK than IMRD-EMIS, which is thought to reflect more affluent groups. This suggests some amount of selection bias and that underlying cohort differences play a role in our findings in each database, a critical consideration for contextualizing our findings and future network studies.

Finally, there were important strengths and limitations of this work. Our comparison of two distinct administrative FIs across 5 distinct CDM databases and two countries revealed critical threats to the external validity of FIs outside the contexts in which they were validated. We were limited to a one-year lookback for US databases, which may have resulted in selection bias because of the inability to track individuals through switching of private plans in Pharmetrics+ and the recency with which *All of Us* began collecting EHR. However, the validity of this approach has been shown; one year is the most used lookback for frailty calculations in US data. Moreover, the validation studies of the VAFI using one and three-year lookback periods demonstrated little difference extending beyond one year, and the eFI has no predefined lookback period (48,49). The VAFI was developed and validated in the US veteran population, suggesting the potential for misclassification. However, the VAFI has been shown to be comparable to other frailty measures validated outside the US veteran population. There was no laboratory measurement data available in three of the datasets. Only IMRD data contained measurements, which made only modest reductions to the proportion of individuals categorized as frail.

## Conclusion

Using existing FIs in network studies of frailty outside of the demographic or geographic region in which they were developed may not be a valid approach to studying international trends in frailty. We speculate that potential threats to the external validity of international applications of frailty include differences in documentation, translation to a CDM, and cultural norms about utilization and practice. It may be necessary to create a frailty index that is not tied to any one database, but rather is validated for use amidst the complexities of translated multi-source health data, such as the OMOP CDM.

Key takeaways:

- FIs appear to be dependent on their development context, limiting their external validity in international applications
- CDMs may alter the validity of FI measures due to details lost in translation from source data to harmonized data.

## Data Availability

Raw Data from the AllofUs Research Program and UK Biobank are available to researchers who enroll in these respective programs. Access to data from the IMRD and Pharmetrics+ databases were licensed to respective institutions and are not publicly available. Aggregated data from this study are available at https://github.com/rbcavanaugh/frailty-comparisons.

## Acknowledgements

The All of Us Research Program is supported by the National Institutes of Health, Office of the Director: Regional Medical Centers, Federally Qualified Health Centers, Data and Research Center, Biobank, The Participant Center, Participant Technology Systems Center, Communications and Engagement, and Community Partners. We gratefully acknowledge *All of Us* participants for their contributions, without whom this research would not have been possible. We also thank the National Institutes of Health’s *All of Us* Research Program for making available the participant data examined in this study.

## Data Availability Statement

Data from the AllofUs Research Program and UK Biobank are available to researchers who enroll in these respective programs. Data from the IMRD and Pharmetrics+ databases were licensed to respective institutions (KI Institute and Northeastern University) and are not publicly available.

## CRediT

Robert Cavanaugh: Methodology, Software, Formal Analysis, Writing – original draft, writing – review & editing, Visualization

Brianne Olivieri-Mui: Conceptualization, Methodology, Writing – original draft, writing – review & editing, supervision

Chelsea Wong: Conceptualization, writing – review & editing

Louisa H. Smith: Conceptualization, Methodology, writing – review & editing

Maytal Bivas-Benita: Conceptualization, Data Curation, writing – review & editing

Pinchas Akiva: Conceptualization, Data Curation, writing – review & editing

Tal El-Hay: Conceptualization, Data Curation, writing – review & editing

Ariela R. Orkaby: Conceptualization, writing – review & editing

Chen Yanover: Conceptualization, Methodology, Formal Analysis, Software, Writing – original draft, writing – review & editing, supervision

## Supplemental Materials

**Supplemental Figure 1.**
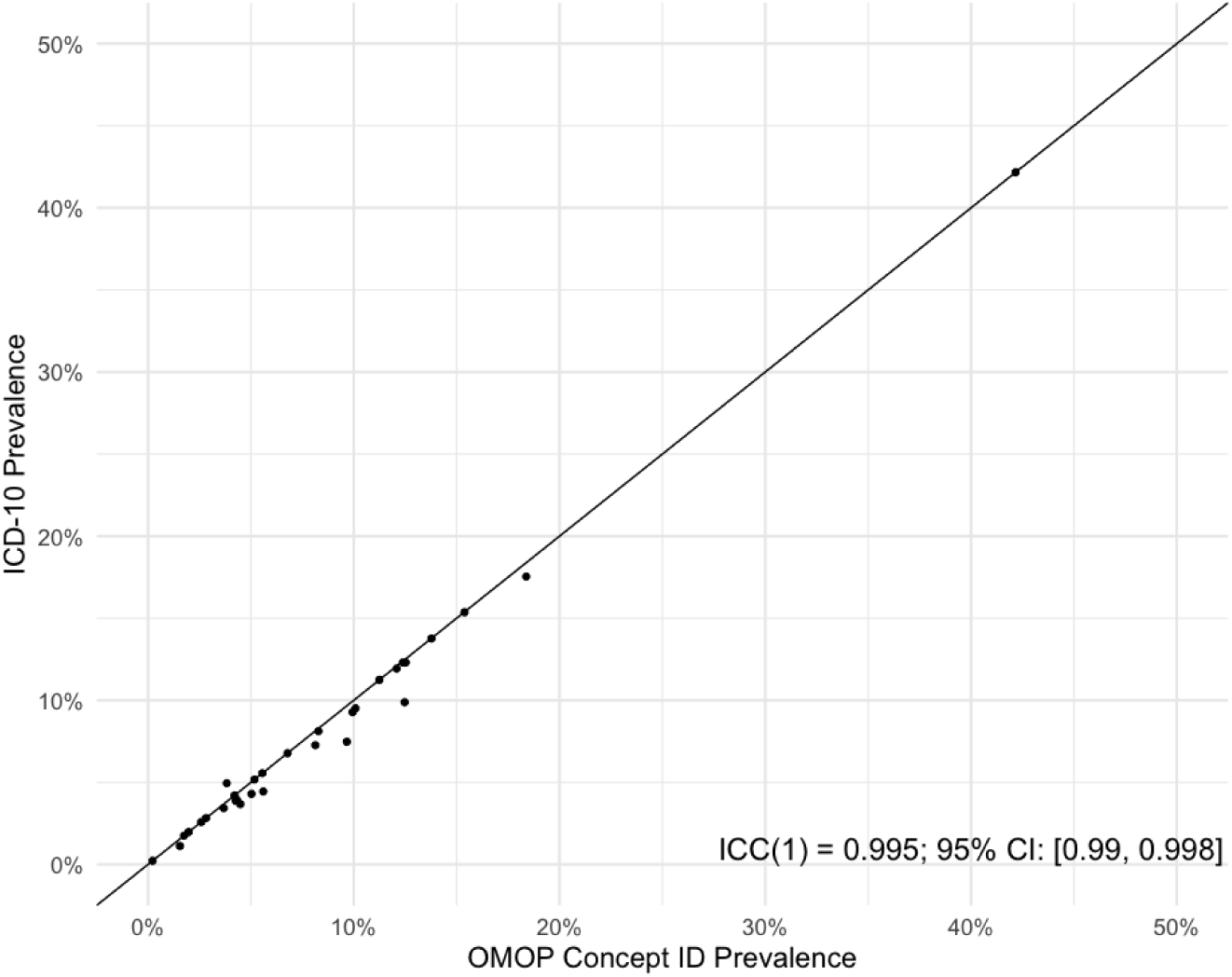
One-way agreement between VAFI deficit prevalence using the OMOP CDM concept codes and the source data ICD-10 codes from Pharmetrics+ to assess the quality of data transformation. *Note: the correlation coefficient demonstrated high correlation (ICC=0.995) between the OMOP CDM transformation and the source coding for deficits for each FI. This implies validity of the data transformation*.

**Supplemental Table 1.** Consort table with inclusion criteria for all datasets.

|  | Pharmetrics | All of Us | IMRD-<br>EMIS | IMRD-<br>THIN | UKBB |
| --- | --- | --- | --- | --- | --- |
| People in data | 34,808,145 | 413,360 | 5,187,327 | 13,209,954 | 502,363 |
| Visit in 2010 or later | 28,076,893 | 287,012 | 2,969,868 | 8,038,288 | 501,472 |
| Sufficient lookback* | 9,016,272 | 214,375 | 1,480,491 | 5,091,067 | 207,202 |
| Age >=40 years | 5,099,557 | 159,721 | 832,455 | 3,036,003 | 207,202 |
| Final sample | 5,099,557 | 159,721 | 832,455 | 3,036,003 | 207,202 |

\* At least 1 year in US databases or 3 years in UK databases

**Supplemental Table 2.** Sensitivity Analysis using 3-year lookback and measurement data.

|  | IMRD-EMIS<br>(N=832,455) |  | IMRD-THIN<br>(N=3,036,003) |  | UK Biobank<br>(N=207,202) |  |
| --- | --- | --- | --- | --- | --- | --- |
| eFI (1y lookback) |  |  |  |  |  |  |
| Frail | 3,886 | (0.5%) | 3,097 | (0.1%) | 283 | (0.1%) |
| Pre-frail | 45,259 | (5.4%) | 95,111 | (3.1%) | 7,061 | (3.4%) |
| Robust | 783,310 | (94.1%) | 2,937,795 | (96.8%) | 199,858 | (96.5%) |
| eFI (1y lookback + measurements) |  |  |  |  |  |  |
| Frail | 5,534 | (0.7%) | 5,643 | (0.2%) |  |  |
| Pre-frail | 55,424 | (6.7%) | 134,887 | (4.4%) |  |  |
| Robust | 771,497 | (92.7%) | 2,895,473 | (95.4%) |  |  |
| eFI (3y lookback) |  |  |  |  |  |  |
| Frail | 18,082 | (2.2%) | 32,416 | (1.1%) | 1,172 | (0.6%) |
| Pre-frail | 90,896 | (10.9%) | 277,392 | (9.1%) | 15,528 | (7.5%) |
| Robust | 723,477 | (86.9%) | 2,726,195 | (89.8%) | 190,502 | (91.9%) |
| eFI (3y lookback + measurements) |  |  |  |  |  |  |
| Frail | 23,775 | (2.9%) | 48,714 | (1.6%) |  |  |
| Pre-frail | 102,599 | (12.3%) | 330,436 | (10.9%) |  |  |
| Robust | 706,081 | (84.8%) | 2,656,853 | (87.5%) |  |  |
| VAFI (1y lookback) |  |  |  |  |  |  |
| Frail | 753 | (0.1%) | 129 | (0.0%) | 69 | (0.0%) |
| Pre-frail | 16,016 | (1.9%) | 8,114 | (0.3%) | 1,541 | (0.7%) |
| Robust | 815,686 | (98.0%) | 3,027,760 | (99.7%) | 205,592 | (99.2%) |
| VAFI (3y lookback) |  |  |  |  |  |  |
| Frail | 4,735 | (0.6%) | 3,073 | (0.1%) | 408 | (0.2%) |
| Pre-frail | 46,344 | (5.6%) | 66,216 | (2.2%) | 5,220 | (2.5%) |
| Robust | 781,376 | (93.9%) | 2,966,714 | (97.7%) | 201,574 | (97.3%) |

## References

1. Kim DH, Rockwood K. Frailty in Older Adults. N. Engl. J. Med. 2024;391(6):538–548.

2. Shrauner W, Lord EM, Nguyen X-MT, et al. Frailty and cardiovascular mortality in more than 3 million US Veterans. Eur. Heart J. 2022;43(8):818–826.

3. Searle SD, Mitnitski A, Gahbauer EA, et al. A standard procedure for creating a frailty index. BMC Geriatr. 2008;8(1):24–24.

4. Rockwood K, Mitnitski A. Frailty in Relation to the Accumulation of Deficits. J. Gerontol. Ser. A. 2007;62(7):722–727.

5. Mitnitski AB, Mogilner AJ, MacKnight C, et al. The mortality rate as a function of accumulated deficits in a frailty index. Mech. Ageing Dev. 2002;123(11):1457–1460.

6. Cheng D, Dumontier C, Sheikh AR, et al. Prognostic value of the veterans affairs frailty index in older patients with non-small cell lung cancer. Cancer Med. 2022;11(15):3009–3022.

7. Gilbert T, Neuburger J, Kraindler J, et al. Development and validation of a Hospital Frailty Risk Score focusing on older people in acute care settings using electronic hospital records: an observational study. Lancet Lond. Engl. 2018;391(10132):1775–1782.

8. Kim DH, Schneeweiss S, Glynn RJ, et al. Measuring Frailty in Medicare Data: Development and Validation of a Claims-Based Frailty Index. J. Gerontol. A. Biol. Sci. Med. Sci. 2018;73(7):980–987.

9. Cheng D, DuMontier C, Yildirim C, et al. Updating and Validating the U.S. Veterans Affairs Frailty Index: Transitioning From ICD-9 to ICD-10. J. Gerontol. A. Biol. Sci. Med. Sci. 2021;76(7):1318–1325.

10. Kochar A, Deo SV, Charest B, et al. Preoperative frailty and adverse outcomes following coronary artery bypass grafting surgery in US veterans. J. Am. Geriatr. Soc. 2023;71(9):2736–2747.

11. Clegg A, Bates C, Young J, et al. Development and validation of an electronic frailty index using routine primary care electronic health record data. Age Ageing. 2016;45(3):353–360.

12. Sison SDM, Shi SM, Kim KM, et al. A crosswalk of commonly used frailty scales. J. Am. Geriatr. Soc. 2023;71(10):3189–3198.

13. Kim DH, Cheslock M, Sison SM, et al. eFrailty: Making frailty assessment accessible to clinicians and researchers. J. Am. Geriatr. Soc. 2024;

14. Voss EA, Makadia R, Matcho A, et al. Feasibility and utility of applications of the common data model to multiple, disparate observational health databases. J. Am. Med. Inform. Assoc. 2015;22(3):553–564.

15. Weiskopf NG, Weng C. Methods and dimensions of electronic health record data quality assessment: enabling reuse for clinical research. J. Am. Med. Inform. Assoc. JAMIA. 2013;20(1):144–151.

16. Schuemie M, Suchard M, Ryan P, et al. FeatureExtraction: Generating Features for a Cohort. 2024;3.12.0. (https://CRAN.R-project.org/package=FeatureExtraction). (Accessed November 11, 2025)

17. Orfila F, Carrasco-Ribelles LA, Abellana R, et al. Validation of an electronic frailty index with electronic health records: eFRAGICAP index. BMC Geriatr. 2022;22(1):404.

18. OHDSI. The Book of OHDSI. OHDSI; 2019.(https://books.google.co.il/books?id=JxpnzQEACAAJ)

19. All of Us Research Program Investigators, Denny JC, Rutter JL, et al. The “All of Us” Research Program. N. Engl. J. Med. 2019;381(7):668–676.

20. Edwards L, Pickett J, Ashcroft DM, et al. UK research data resources based on primary care electronic health records: review and summary for potential users. BJGP Open. 2023;7(3):BJGPO.2023.0057.

21. Candore G, Hedenmalm K, Slattery J, et al. Can We Rely on Results From IQVIA Medical Research Data UK Converted to the Observational Medical Outcome Partnership Common Data Model?: A Validation Study Based on Prescribing Codeine in Children. Clin. Pharmacol. Ther. 2020;107(4):915–925.

22. Sudlow C, Gallacher J, Allen N, et al. UK Biobank: An Open Access Resource for Identifying the Causes of a Wide Range of Complex Diseases of Middle and Old Age. PLOS Med. 2015;12(3):e1001779.

23. Allen N, Sudlow C, Downey P, et al. UK Biobank: Current status and what it means for epidemiology. Health Policy Technol. 2012;1(3):123–126.

24. Ambagtsheer RC, Beilby J, Dabravolskaj J, et al. Application of an electronic Frailty Index in Australian primary care: data quality and feasibility assessment. Aging Clin. Exp. Res. 2019;31(5):653–660.

25. Morales DR, Guthrie B, Downes TJ, et al. Applicability of the electronic frailty index in younger and older adults in England: a population-based cohort study. Lancet Healthy Longev. 2025;6(8):100752.

26. Kojima G, Iliffe S, Walters K. Frailty index as a predictor of mortality: a systematic review and meta-analysis. Age Ageing. 2018;47(2):193–200.

27. DuMontier C, Hennis R, Yilidirim C, et al. Construct validity of the electronic Veterans Affairs Frailty Index against clinician frailty assessment. J. Am. Geriatr. Soc. 2023;71(12):3857–3864.

28. Kochar B, Cheng D, Lehto H, et al. Application of an Electronic Frailty Index to Identify HighlRisk Older Adults Using Electronic Health Record Data. J. Am. Geriatr. Soc. 2025;73(5):1491–1497.

29. Orkaby AR, Nussbaum L, Ho Y-L, et al. The Burden of Frailty Among US Veterans and Its Association With Mortality, 2002-2012. J. Gerontol. A. Biol. Sci. Med. Sci. 2019;74(8):1257–1264.

30. Clegg A, Bates C, Young J, et al. Development and validation of an electronic frailty index using routine primary care electronic health record data. Age Ageing. 2016;45(3):353–360.

31. Elhussein L, Burn E, Delmestri A, et al. Quantifying polypharmacy in elderly people: a comparison between source and mapped data in the UK Clinical Practice Research Datalink GOLD. In: Abstract presentations. Online: 2021 (Accessed September 5, 2024)(https://www.ohdsi.org/2021-global-symposium-showcase-36/). (Accessed September 5, 2024)

32. Wickham H, Girlich M, Ruiz E. dbplyr: A “dplyr” Back End for Databases. 2017;2.5.1. (https://CRAN.R-project.org/package=dbplyr). (Accessed November 11, 2025)

33. Shi SM, Olivieri-Mui B, Park CM, et al. Frailty in Medicare Advantage Beneficiaries and Traditional Medicare Beneficiaries. JAMA Netw. Open. 2024;7(8):e2431067.

34. Sinclair DR, Maharani A, Chandola T, et al. Frailty among Older Adults and Its Distribution in England. J. Frailty Aging. 2022;11(2):163–168.

35. Kent S, Burn E, Dawoud D, et al. Common Problems, Common Data Model Solutions: Evidence Generation for Health Technology Assessment. Pharmacoeconomics. 2021;39(3):275–285.

36. Jacob Z. A Comparative Analysis of the US and UK Health Care Systems – Michigan Journal of Economics. Mich. J. Econ. [electronic article]. 2023;Online. (https://sites.lsa.umich.edu/mje/2023/05/26/a-comparative-analysis-of-the-us-and-uk-health-care-systems/). (Accessed August 13, 2025)

37. Jia L, Meng Q, Scott A, et al. Payment methods for healthcare providers working in outpatient healthcare settings. Cochrane Database Syst. Rev. 2021;1(1):CD011865.

38. Petrou P, Samoutis G, Lionis C. Single-payer or a multipayer health system: a systematic literature review. Public Health. 2018;163:141–152.

39. Tsou AY, Lehmann CU, Michel J, et al. Safe Practices for Copy and Paste in the EHR. Appl. Clin. Inform. 2017;8(1):12–34.

40. Documenting in patient clinical recordsl: CPPE. (https://www.cppe.ac.uk/programmes/l/clinical-e-02). (Accessed August 13, 2025)

41. All of Us Research Program Investigators, Denny JC, Rutter JL, et al. The “All of Us” Research Program. N. Engl. J. Med. 2019;381(7):668–676.

42. van Alten S, Domingue BW, Faul J, et al. Reweighting UK Biobank corrects for pervasive selection bias due to volunteering. Int. J. Epidemiol. 2024;53(3):dyae054.

43. Kambara MS, Sharma S, Spouge JL, et al. Increasing Representativeness in the All of Us Cohort Using Inverse Probability Weighting. medRxiv. 2024;2024.10.02.24314774.

44. Gordon EH, Peel NM, Samanta M, et al. Sex differences in frailty: A systematic review and meta-analysis. Exp. Gerontol. 2017;89:30–40.

45. Haynes K, Bilker WB, TenHave TR, et al. Temporal and Within Practice Variability in The Health Improvement Network. Pharmacoepidemiol. Drug Saf. 2011;20(9):948–955.

46. Denburg MR, Haynes K, Shults J, et al. Validation of The Health Improvement Network (THIN) database for epidemiologic studies of chronic kidney disease. Pharmacoepidemiol. Drug Saf. 2011;20(11):1138–1149.

47. Lewis JD, Schinnar R, Bilker WB, et al. Validation studies of the health improvement network (THIN) database for pharmacoepidemiology research. Pharmacoepidemiol. Drug Saf. 2007;16(4):393–401.

48. Correction to: Comparison of Claims-Based Frailty Indices in U.S. Veterans 65 and Older for Prediction of Long-Term Institutionalization and Mortality. J. Gerontol. Ser. A. 2024;79(2):glad246.

49. Orkaby AR, Huan T, Intrator O, et al. Comparison of Claims-Based Frailty Indices in U.S. Veterans 65 and Older for Prediction of Long-Term Institutionalization and Mortality. J. Gerontol. Ser. A. 2023;78(11):2136–2144.

